# Applications of fluorescence-guided surgery across multiple tumor types using a near-infrared labeled EGFR antibody

**DOI:** 10.1101/2021.10.15.21265068

**Authors:** Quan Zhou, Nynke S. van den Berg, Wenying Kang, Jacqueline Pei, Naoki Nishio, Stan van Keulen, Myrthe A. Engelen, Yu-Jin Lee, Marisa Hom, Johana C. M. Vega Leonel, Zachary Hart, Hannes Vogel, Romain Cayrol, Brock A. Martin, Mark Roesner, Glenn Shields, Natalie Lui, Melanie Hayden Gephart, Roan C. Raymundo, Grace Yi, Monica Granucci, Gerald A. Grant, Gordon Li, Eben L. Rosenthal

**Affiliations:** Department of Otolaryngology, Stanford University School of Medicine, Stanford, CA, USA; Department of Neurosurgery, Stanford University School of Medicine, Stanford, CA, USA; Department of Otorhinolaryngology, Nagoya University Graduate School of Medicine, Nagoya, Japan; Department of Oral and Maxillofacial Surgery and Oral Pathology, Amsterdam UMC-location VUMC/Academic Centre for Dentistry Amsterdam (ACTA), Amsterdam, The Netherlands; Department of Mechanical Engineering, Delft University of Technology, Delft, Netherlands; Department of Pathology, Stanford University, Stanford, CA, USA; Stanford Health Care, Stanford University Medical Center, Stanford, CA, USA; Department of Cardiothoracic Surgery, Stanford University Medical Center, Stanford, CA, USA; Cancer Clinical Trials Office, Stanford University School of Medicine, Stanford, CA, USA

**Keywords:** Near-infrared fluorescence imaging, panitumumab-IRDye800, epidermal growth factor receptor, clinical trial, high-grade glioma, head and neck squamous cell carcinoma, lung adenocarcinoma

## Abstract

**Background:** As receptor-ligand based strategies emerge for surgical imaging, the relative importance of receptor expression in different tumor types is unknown. Near-infrared (NIR) labeled epidermal growth factor receptor (EGFR) antibody, panitumumab-IRDye800, was evaluated across three cancers to demonstrate its clinical utilities and a holistic analysis framework.

**Methods:** Thirty-one patients diagnosed with high-grade glioma (HGG, n=5, NCT03510208), head and neck squamous cell carcinoma (HNSCC, n=23, NCT02415881) or lung adenocarcinoma (LAC, n=3, NCT03582124) received systemic administration of 50 mg panitumumab-IRDye800 days prior to surgery. Intraoperative NIR laparoscopic or open-field images of the surgical field were acquired and tissue mimicking phantoms were constructed to identify optimal imaging conditions. Margin distance was correlated to fluorescence on resected specimen surface. Panitumumab-IRDye800 distribution was registered to histology in fixed tissue sections. Immunohistochemistry characterized EGFR expression.

**Results:** Intraoperative NIR imaging enhanced tumor contrast against surrounding healthy tissue by 5.2-fold, 3.4-fold and 1.4-fold in HGG, HNSCC and LAC, respectively. Imaging quality was optimal at the lowest gain possible under ambient light. Ex vivo NIR fluorescence identified 78 – 97% of at-risk resection margins, with 72 – 92% sensitivity and 67 – 96% specificity for tumor in fixed tissue sections. Intratumoral panitumumab-IRDye800 concentration correlated with total tumoral EGFR expression (HGG > HNSCC > LAC) and delivery barrier. Cellular EGFR expression (80%) and tumor cell density (3000 cells/mm2) was highest in HGG.

**Conclusions:** In multiple tumor types, EGFR-targeting in fluorescence-guided surgery translated to enhanced macroscopic tumor contrast and successful margin assessment despite disparate tumor cell density and heterogeneous delivery of pantimumab-IRDye800.

**Translational relevance:** As receptor-ligand based strategies emerge for surgical imaging, the relative importance of receptor expression in different tumor types remains unexamined. Near-infrared labeled epidermal growth factor receptor (EGFR) antibody was evaluated across three cancers to demonstrate its clinical utilities and a holistic analysis framework. Targeted fluorescence imaging was performed in patients infused with a consistent dose of panitumumab-IRDye800 using the same fluorescence imaging devices. In high-grade glioma, head and neck squamous cell carcinoma and lung adenocarcinoma, EGFR-targeting in fluorescence-guided surgery translated to enhanced macroscopic tumor contrast and successful margin assessment. While total EGFR expression and delivery barrier corresponded to overall panitumumab-IRDye800 distribution in tissue, tumor depth and operating room lighting can have greater influence over the intraoperative fluorescence contrast at specific tissue surface locations of a certain patient. These factors can facilitate selecting appropriate molecular target and tumor qualities to streamline translation across institutions and regulatory approval of new imaging probes.

## Introduction

Surgery remains the most common first-line treatment for cancer. In the past decade, tumor-specific fluorescent tracers have demonstrated safety [1–3] and ability to enhance tumor visualization [4–13] in cancers of the head and neck, brain, breast, ovary, pancreas, kidney and skin. However, previous studies typically reported findings on one cancer type or performed fluorescence imaging using different imaging instruments and analysis methods. While receptor-ligand based intraoperative imaging probes targeting folate receptor (OTL38 [14, 15]), carcinoembryonic antigen (SGM-101 [16]) and prostate specific membrane antigen [17–19] achieved early successes in imaging ovarian, colorectal and prostate cancers, the underlying mechanism of how microscopic tumor specific receptor overexpression translates to macroscopic fluorescence contrast remains unexamined. As more receptor-based imaging probes enter phase II and III clinical trials, there is a need to compare the performance of probes across different tumor types and identify factors that may help design an optimal imaging strategy.

For early phase studies with dose escalation structures, fluorescence imaging between different dose cohorts can be naturally compared within the same analysis framework following recommendations proposed for reporting emerging optical imaging agents [20], as illustrated in breast cancer [21]. Yet so far no consensus has been reached on evaluating a fluorescence imaging probe for one cancer type against others in a clinical setting. In this study, we compared open-field imaging performance, pathological margin assessment and tissue distribution of a fluorescent antibody in high-grade glioma (HGG), head and neck squamous cell carcinoma (HNSCC) and lung adenocarcinoma (LAC). Imaging results were correlated with target expression (epidermal growth factor receptor, EGFR) and tumor cell density. Patients received a single dose of near-infrared (NIR) labeled EGFR antibody, panitumumab-IRDye800, in our analytical pipeline encompassing multiple tumor types acquired with the same four fluorescence imaging instruments to demonstrate a framework that might improve clinical translation and regulatory approval of emerging molecular targeted imaging probes.

## Results

### Summary of study design

Three single center, open-label, phase I/II studies were performed in 31 patients with HGG (n = 5), HNSCC (n = 23) and LAC (n = 3) undergoing surgical resection of the primary tumor. Patients received an infusion with panitumumab-IRDye800CW 1–5 days prior to surgery at a single dose of 50 mg. On the day of surgery, intraoperative NIR fluorescence imaging was performed to assess tumor and wound bed in *vivo* using an open-field device (with either a handheld or laparoscopic camera). After surgical resection, the resected specimens were imaged in a closed-field fluorescence device and cut into segments exposing tumor cross sections. Formalin-fixed and paraffin-embedded (FFPE) tissue blocks and corresponding tissue sections were imaged in a flatbed fluorescence scanner and examined under fluorescence microscope. In collaboration with three board-certified pathologists (H.V., R.C. and B.A.M), molecular imaging results were correlated to immunohistochemical (IHC) and pathological (hematoxylin and eosin, H&E) findings, as illustrated in our proposed tissue processing and imaging workflow in **Fig. 1**.

**Figure 1.**
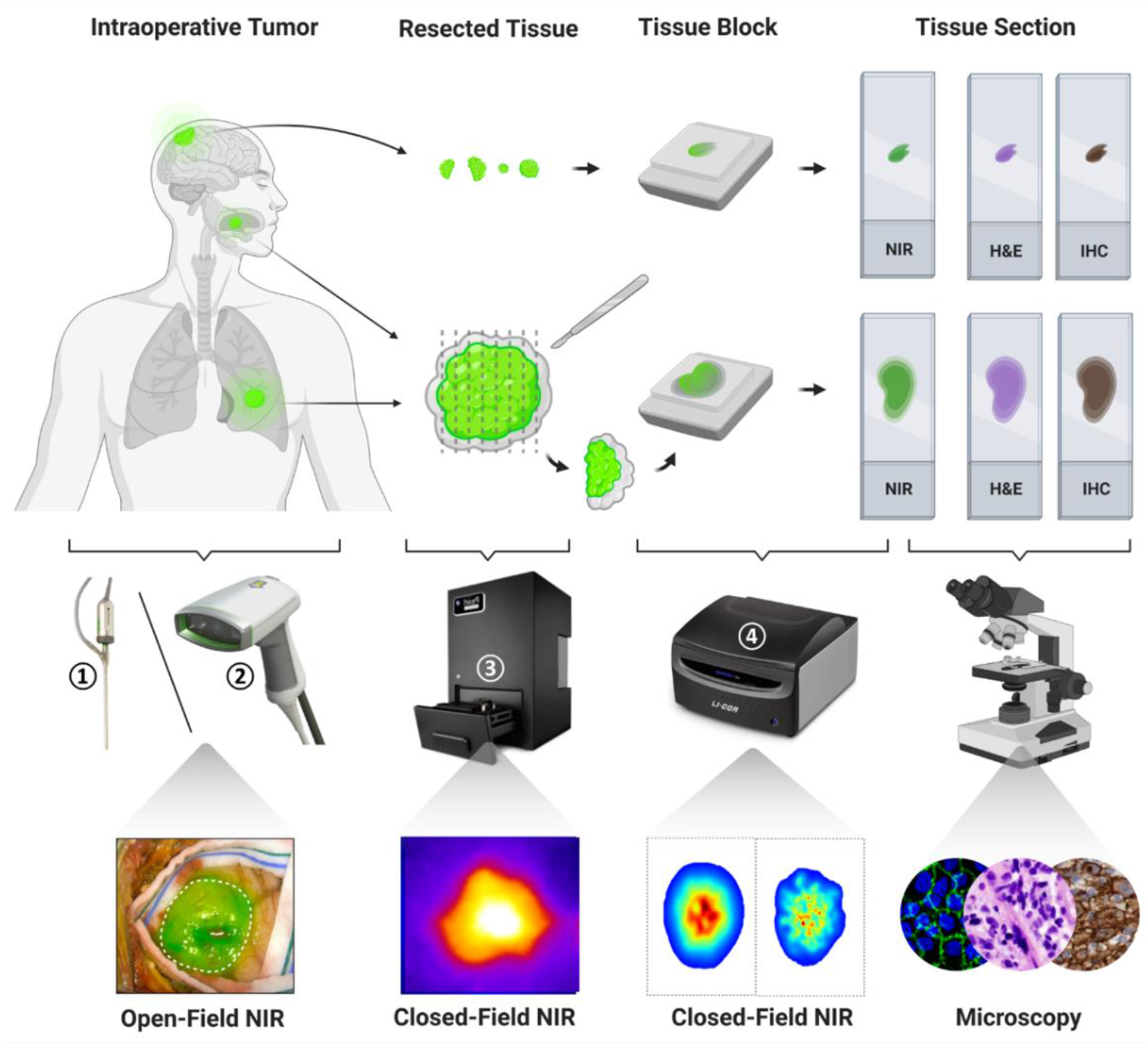
Tissue processing and imaging workflow. Intraoperative near-infrared (NIR) fluorescence imaging was performed in patients undergoing cancer resection surgery who were infused with panitumumab-IRDye800CW prior to surgery. NIR fluorescence images were taken of resected fresh tissue before being fixed and paraffin embedded in tissue blocks (after large solid tumors were cross-sectioned into 5mm segments). Macroscopic and microscopic NIR fluorescence images were collected of fixed tissue and corresponding histopathological stainings post surgery. Open-field and closed-field NIR fluorescence imaging instruments (vendors): ① laparoscope and ② portable handheld imager of the SPY imaging platform (Novadaq); ③ Pearl Trilogy (Li-COR Biosciences); ④ Odyssey CLx (Li-COR Biosciences). Images reproduced with permission from Stryker, and LI-COR, Inc.

### Patient characteristics

The genders were unevenly split (M/F = 0.72) among study participants, 55% of whom above 65 years old, **Table 1**. A majority of Caucasian (84%) patients were mixed with four (13%) Asians and one (3%) of unknown race. Tumor size was similar among trials measuring 4.1 ± 0.4 cm (*P* = 0.35), while resected tissue size varied significantly between diffuse HGGs removed in pieces and solid tumors resected en bloc (16% ± 4% vs. 184% ± 20% of the tumor size, *P* = 0.0002), **Supplementary Fig. S1**. All patients received 50 mg panitumumab-IRDye800, or a median weight-adjusted dose of 0.7 mg/kg (range: 0.4 - 1.2 mg/kg). On average, 38 hours (range: 14 - 90 hours) separated infusion and first incision when the mean plasma concentration measured 5.5 mg/L (range: 0.8 - 17.3 mg/L). No significant difference was found between trials in demographic features, weight-adjusted tracer dose, plasma probe concentration and imaging window. No infusion reactions or dose limiting toxicity events occurred up to 30 days following surgery, **Supplementary Table S1**, and none of the 58 adverse events (44 grade I, 11 grade II and three grade III) were serious or attributed to panitumumab-IRDye800.

**Table. 1.** Patient characteristics.

|  | **HGG**<br>(n = 5) | **HNSCC**<br>(n = 23) | **LAC**<br>(n = 3) | **Total**<br>(n = 31) | **P value** |
| --- | --- | --- | --- | --- | --- |
| Age, y (median/range) | 62 (42-72) | 67 (44-82) | 71 (67-71) | 67 (42-82) | 0.41<sup>a</sup> |
| Gender, Male (%) | 2 (40%) | 10 (43%) | 1 (33%) | 13 (42%) | 0.94<sup>b</sup> |
| Race |  |  |  |  | 0.31<sup>b</sup> |
| Asian | 1 (20%) | 2 (9%) | 1 (33%) | 4 (13%) |  |
| White | 4 (80%) | 20 (87%) | 2 (67%) | 26 (84%) |  |
| Unknown/Not reported | 0 (0%) | 1 (4%) | 0 (0%) | 1 (3%) |  |
| Tumor size, cm (median/range) | 5.0 (3.5-6.1) | 2.8 (1.0-9.0) | 2.3 (1.9-3.5) | 3.7 (1.0-9.0) | 0.35<sup>a</sup> |
| Pan800 dose, mg/kg (mean ± SD) | 0.8 ± 0.3 | 0.8 ± 0.2 | 0.6 ± 0.3 | 0.7 ± 0.2 | 0.84<sup>a</sup> |
| Pan800 DOS plasma conc., mg/L (mean± SD) | 6.9 ± 3.5 | 5.5 ± 4.2 | 3.9 ± 3.7 | 5.5 ± 4.0 | 0.58<sup>a</sup> |
| Imaging window, days, (median/range) | 1.8 (0.6-2.9) | 1.8 (0.8-3.8) | 1.7 (0.9-1.8) | 1.8 (0.6-3.8) | 0.95<sup>a</sup> |

<sup>a</sup> One-way AVOVA; <sup>b</sup> Pearson's chi-squared test. \*Statistically significant at p < 0.05
HGG: high-grade glioma; HNSCC: head-and-neck squamous cell carcinoma; LAC: lung adenocarcinoma; DOS: day of surgery.

### Intraoperative tumor visualization

Intraoperative fluorescence was diffuse in LAC compared to the strong signal in HGG and HNSCC that allowed distinct separation of disease tissue from normal areas based on histological confirmation, with notable heterogeneity in HGG, **Fig. 2A**. Minimal fluorescence remained in the wound beds of HNSCC and LAC, while fluorescent residual HGG involving eloquent cortex located beyond MRI contrast-enhancing margin was not removed in the resection cavity, **Supplementary Fig. S2**. NIR imaging enhanced tumor contrasts relative to white light illumination by 5.2, 3.4 and 1.4 folds (*P* = 0.0006, < 0.0001, and = 0.03, respectively) for HGG, HNSCC and LAC, **Fig. 2B**. TBR correlated with resected tumor size (*P* = 0.007), **Supplementary Fig. S3**. Fluorescence contrasts dropped below 1.0 in the wound beds.

**Figure 2.**
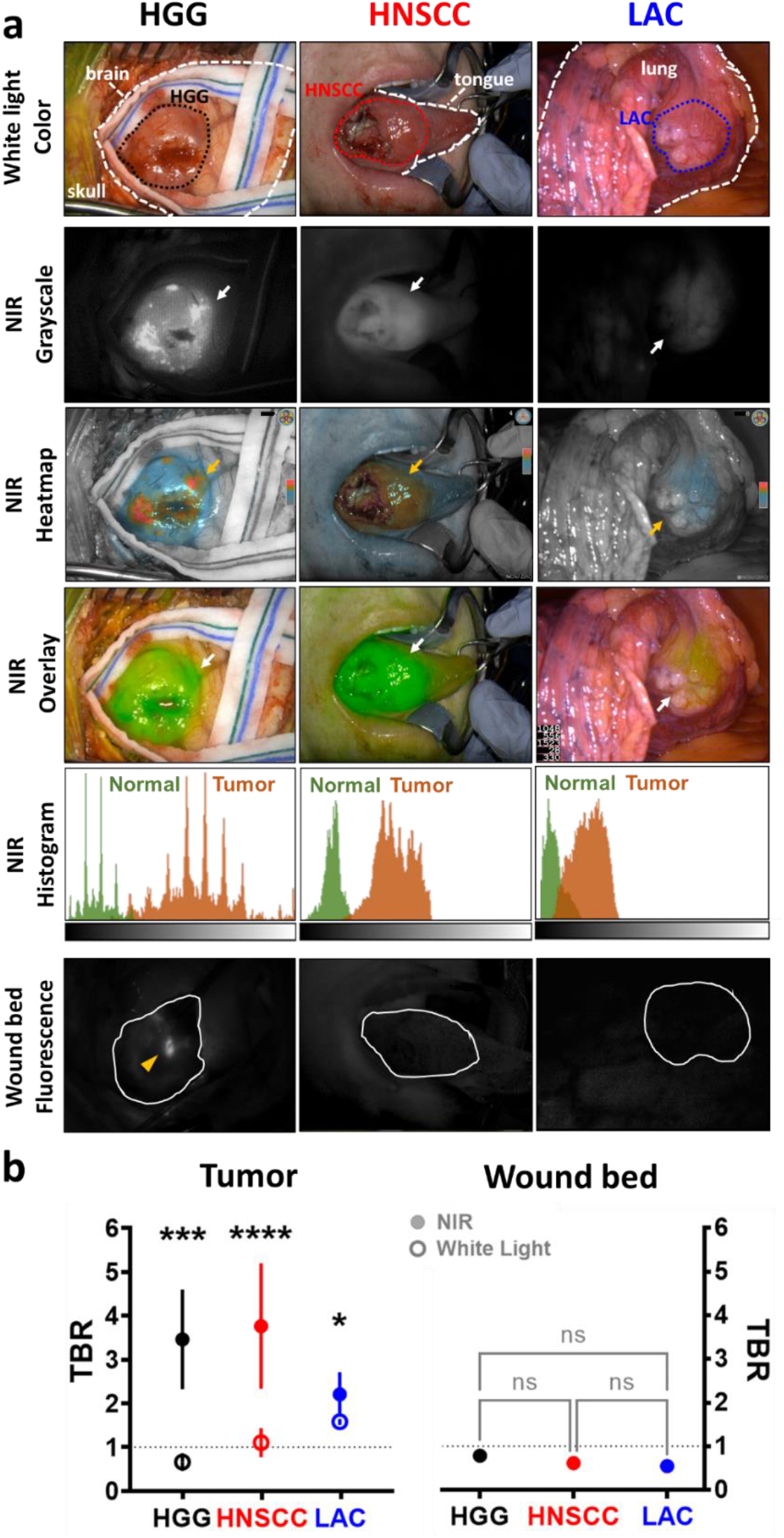
Intraoperative NIR fluorescence imaging enhances tumor contrast *in vivo*. (**a**) Representative annotated (*dashed lines*) white light photographs of exposed tumors (*dotted outlines*) in the surgical field, and corresponding NIR fluorescence images (in grayscale, heatmap and overlay modes) with histograms of fluorescence intensity distribution. NIR fluorescence images were acquired in wound beds (*solid outlines*) after tumor resection. HGG: high-grade glioma; HNSCC: head and neck squamous cell carcinoma; LAC: lung adenocarcinoma. *Arrows*: positive NIR fluorescence signal; *Arrowhead*: residual tumor; Histogram *X-axis*: pixel fluorescence intensity (range: 0 – 255); Histogram *Y-axis*: pixel count (range: 0 – 5000). (**b**) Intraoperative tumor contrasts (target-to-background ratio, TBR) under white light and NIR illumination, versus NIR imaging contrast in wound beds. \*\*\*\**P* < 0.0001, *** *P* = 0.0006, and * *P*= 0.03.

Per workflow requirements, ambient lighting was always present in the operating room during intraoperative NIR imaging. The open-field imaging device had limited sensitivity and dynamic range over tissue-mimicking phantoms containing panitumumab-IRDye800, which could be readily distinguished from each other in a closed-field device (Pearl Trilogy Imaging System, LI-COR Biosciences) that eliminated ambient light, **Supplementary Fig. S4**. MFIs below 0.005 (*yellow circle*) were indistinguishable with open-field imaging at the lowest gain, and MFIs above 0.1 (*red circle*) resulted in saturation. Detection sensitivity was improved (MFI = 0.002, *blue circle*) with higher gain, at the cost of lower saturation threshold (MFI = 0.04, *pink circle*). Operating room lights gave a false positive signal and overhead lights saturated the images in control phantoms, indicating NIR interference from these light sources. The lowest gain was desirable for imaging the majority of HNSCC cases whose MFI interquartile range (IQR) reached 0.08 – 0.23, **Fig. 2F**. In contrast, medium to high gains were often necessary for adequate detection of dim signal in LAC (IQR: 0.02 – 0.11). Low and high gains were adopted for detection of exposed tumor and residual HGG fragments in the wound bed, respectively, as strong fluorescence in en bloc HGG was on par with HNSCC while reduced MFIs (IQR: 0.01 – 0.05) were measured in small resected pieces (7.4 ± 1.2 mm).

### Margin assessment with fluorescence

Closed-field fluorescence images (Pearl Trilogy Imaging System, LI-COR Biosciences) of resected tumor specimens were acquired to identify potentially positive or close margin, **Fig. 3A**. Infiltrative (*dotted outline*) and solid (*solid outline*) tumors were delineated on histology staining of tissue sections sampled across fluorescence intensity peaks, **Fig. 3B**. The HGG cell density decreased beyond the infiltrating edge and distances from tissue surface to tumor margin were inversely correlated with TBR of fluorescence on specimen surface, **Fig. 3C**. Positive and close margins can be captured above the TBR value at 5 mm margin size on fitted regression curves at 97% and 78% detection rates for HNSCC and LAC, respectively, while 93% HGG infiltrative edges with ≥ 50% tumor cell density were detected.

**Figure 3.**
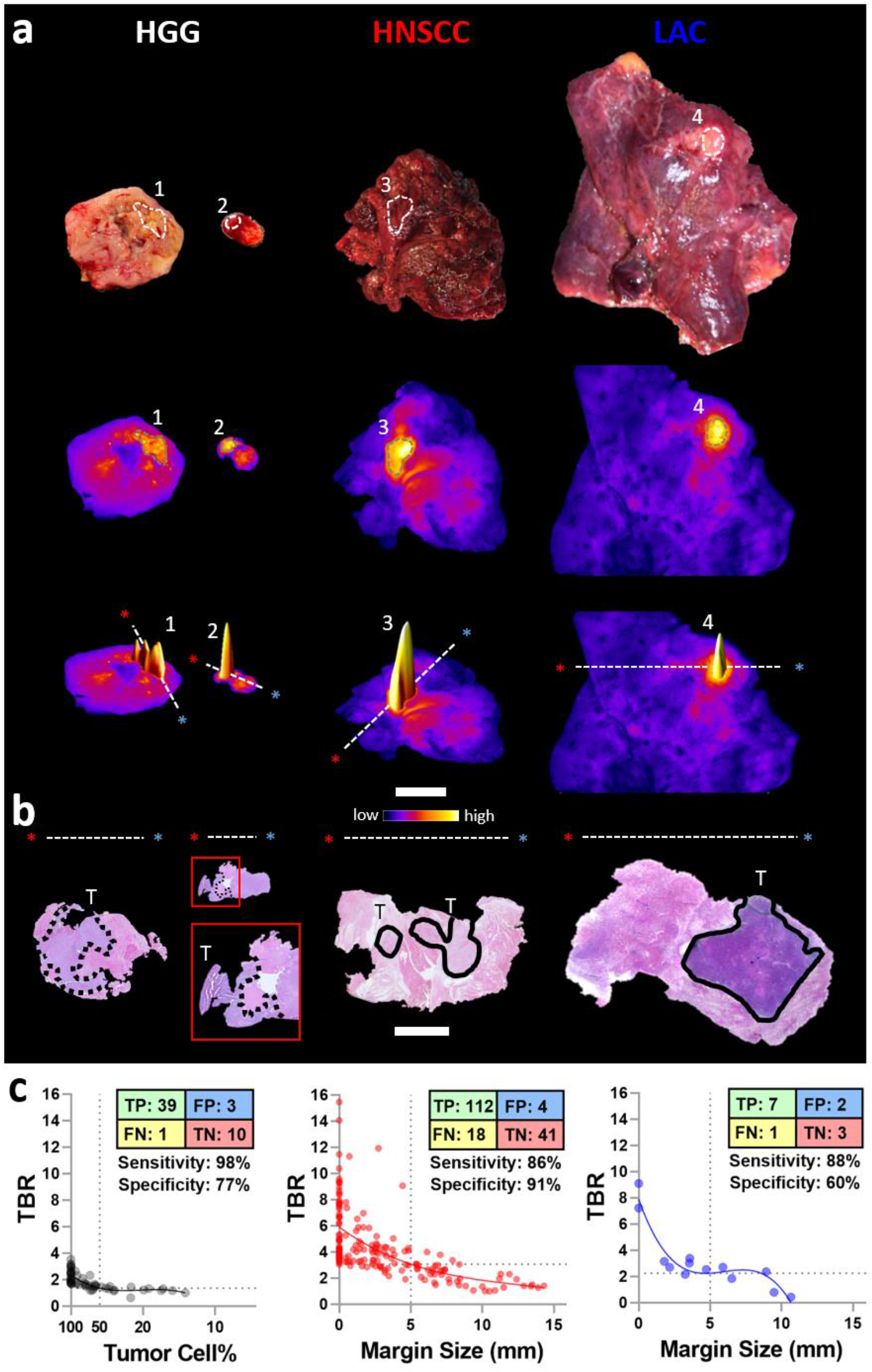
Macroscopic closed-field NIR imaging identifies positive and close margins in resected tissue. (**a**) Representative intraoperative photographs and fluorescence images of resected tissue specimens. Areas 1 – 4 identify the highest fluorescence intensity on each sample. Scale bar = 2 cm. The *dashed lines* and *asterisks* (*red & blue*) indicate the orientation in which (**b**) histology (H&E) slides with infiltrative (*dotted outlines*) and solid (*solid outlines*) tumors (T) were sectioned. Scale bar = 1 cm. (**c**) NIR intensity were plotted against HGG cell percentage and margin size (HNSCC and LAC), respectively. TP: true positive; FP: false positive; TN: true negative; FN: false negative.

### Intratumoral distribution of fluorescent antibody

Microscopic NIR images of FFPE tissue blocks and sections exposing tumor interior confirmed intratumoral distribution and cellular delivery of panitumumab-IRDye800, **Fig. 4A**. Differences in tumor fluorescence converged from 244 to 21 folds as variance in tissue thickness reduced from centimeters in fresh tissue to < 1 μm in tissue sections, **Supplementary Fig. S5**, where NIR signals measured by two closed-field devices were correlated, **Supplementary Fig. S6**. Fluorescence heterogeneity in HGG were more pronounced than in LAC (*P* = 0.02, **Fig. 4B**) with similar tumoral fluorescence contrast, **Fig. 4C**. Fluorescence in tissue sections can effectively distinguish tumor against normal tissue (AUC ≥ 0.85, **Fig. 4D**) in HNSCC, LAC and HCC, consistent with other diagnostic performance parameters, **Supplementary Table S2**. Panitumumab-IRDye800 concentration (inferred from corresponding fluorescence, **Supplementary Fig. S7**) were higher inside tumoral outlines of HGG (3.9 vs 1.6 ng/mg, *P* < 0.0001), HNSCC (8.1 vs 3.9 ng/mg, *P* < 0.0001) and LAC (6.3 vs 4.5 ng/mg, *P* = 0.0006) relative to healthy adjacent tissue, **Fig. 4E**. Further delineation into finer histological structures revealed greater probe distribution in microscopic LAC tumor nodules relative to macroscopic tumoral area, indicating substantial stromal presence inside LAC. Likely due to its EGFR expression, head and neck mucus exhibited distinctly high probe uptake among non-tumoral areas including normal (muscle, lung and brain) tissue, lymph node, stroma, fat, and necrosis.

**Figure 4.**
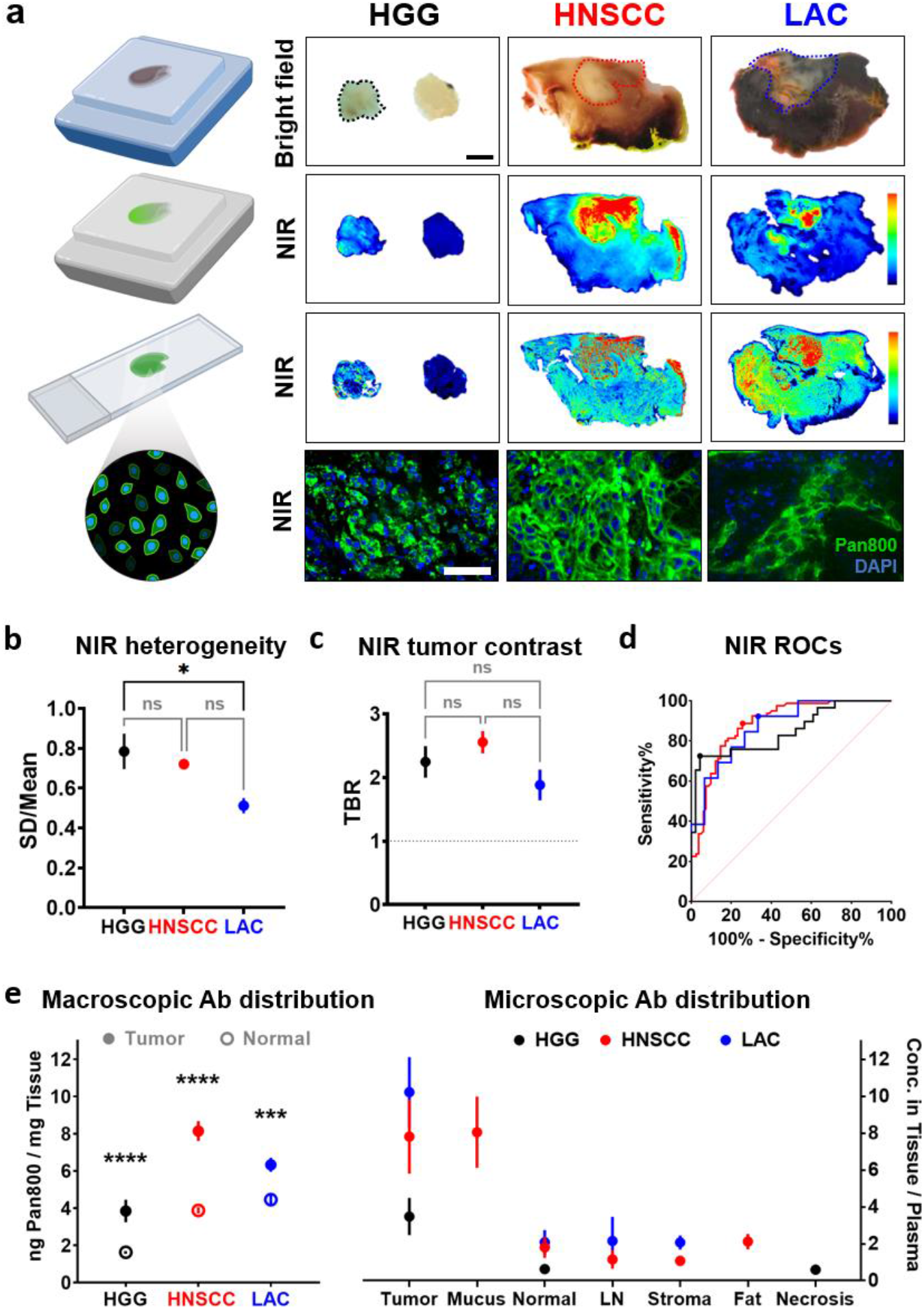
Microscopic closed-field NIR imaging reveals intratumoral distribution and cellular delivery of panitumumab-IRDye800. (**a**) Representative bright field photographs (scale bar = 5 mm; *dotted outlines*: tumor) and fluorescence images (scale bar = 50 μm) of fixed tissue blocks and sections. (**b**) Tumoral fluorescence heterogeneity and (**c**) contrast on tissue sections. * *P* = 0.02. (**d**) ROC curves and performance characteristics of NIR fluorescence for tumor detection. (**e**) Macroscopic and microscopic distribution of panitumumab-IRDye800 in histological tissue types. **** *P* < 0.0001, *** *P* = 0.0006.

### Target validation and tumor cell density

EGFR immunoreactivity heatmaps and histograms illustrated its heterogeneous expression in tumors, which were registered with histology by H&E staining, **Fig. 5A**. This biomarker had greater fidelity for tumor presence (AUCs ≥ 0.9) in HNSCC and HGG than LAC (AUC = 0.82), among other diagnostic performance parameters, **Supplementary Table S3**. Non-specific delivery to peritumoral EGFR negative regions, however, was observed in normal head and neck as well as lung tissue (**Fig. 4A & 5A**). Microscopy revealed intratumoral EGFR staining patterns, contrasting diffuse HGG cells without delineable margin against demarcated focal clusters of HNSCC and LAC tumor cells dispersed among EGFR^−^ stroma and fibroblast. Total tumoral EGFR expression (percentage of positive pixels within the tumor area, *dotted outlines*) was significantly elevated from that of normal tissue in HNSCC (62% vs. 11%), HGG (89% vs. 46%) and LAC (41% vs. 15%), **Fig. 5B**. Higher total tumoral EGFR expression translated to greater panitumumab-IRDye800 concentration in tumors with the notable exception of HGG (**Fig. 4C & 5B**), indicating delivery barrier, **Supplementary Fig. S8**. Cellular EGFR level was particularly high in HGG (80%), but similar between HNSCC and LAC (64% and 60%, respectively). EGFR+ tumor cells (*colored dots*, **Fig. 5C**) were dense in HGG (3000 cells/mm^2^) and HNSCC (2100 cells/mm^2^), but sporadic in LAC (1300 cells/mm^2^) with fewer than 10 cells occupying over 50% tumor areas, **Fig. 5D & Supplementary Fig. S9**. EGFR immunoreactivity thus depended on magnification powers and specific intratumoral locations examined, **Supplementary Fig. S10**. These observations indicate that EGFR expression is not a scale-free characteristic, but rather the interplay of expression within individual tumor cells, tumor cell density (percentage of tumor to stromal tissue) and distribution pattern within the macroscopically identified tumor area.

**Figure 5.**
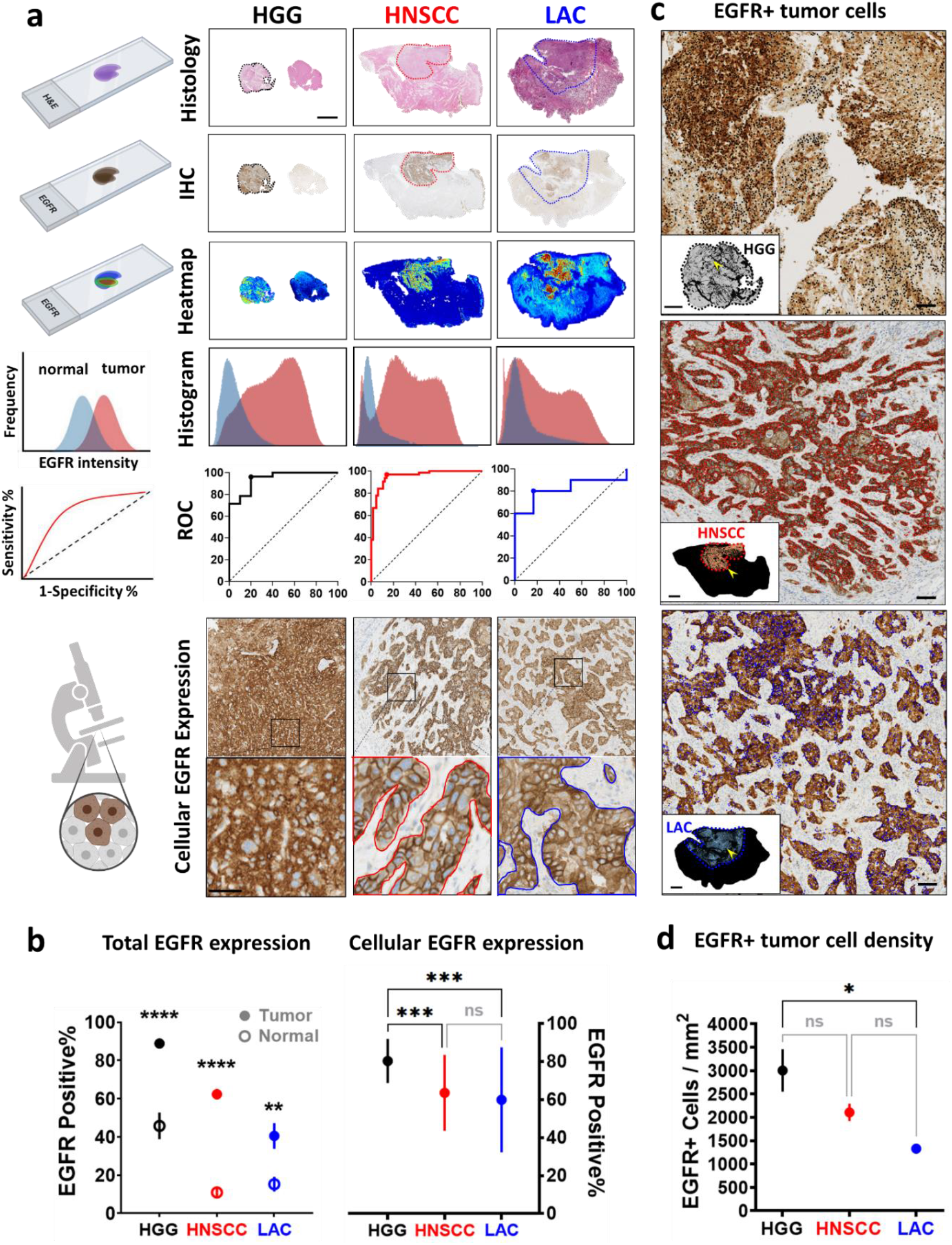
Target validation and tumor cell density. (**a**) Macroscopic tumor areas (scale bar = 5 mm; *dotted outlines*) were registered between histological (H&E) and EGFR immunohistochemical (IHC) stainings on tissue sections with corresponding EGFR expression heatmap and histograms. EGFR expression as a biomarker for different tumor types were plotted as ROC curves. Microscopic tumor nodules (scale bar = 50 μm; *solid outlines*) were examined at subcellular resolution. (**b**) Percentage of pixels with positive EGFR expression in total tumoral and normal areas (**** *P* < 0.0001, ** *P* = 0.003 by multiple t-test), as well as in individual tumor cells (*** *P* = 0.0005, HGG vs HNSCC; *** *P* = 0.001, HGG vs. LAC by ANOVA). (**c**) Each EGFR+ tumor cell is identified with a colored dot (*black*: HGG; *red*: HNSCC; *blue*: LAC; scale bars = 200 μm) within tumor areas. *Insets*: overall distribution of EGFR+ tumor cells on whole tissue sections (scale bar = 2 mm (HGG); scale bars = 2 cm (HNSCC and LAC)); *Arrowheads*: location of high magnification microscopic views. (**d**) EGFR positive tumor cell density. * *P* = 0.025 by ANOVA.

## Discussion

We demonstrated that fluorescence imaging of a NIR-labeled EGFR antibody, panitumumab-IRDye800, can be incorporated into conventional clinical workflow to facilitate intraoperative tumor visualization and margin assessment. In a reductionist and comparative approach to isolate factors that influence intraoperative imaging performance, we went from *in vivo* open-field to *ex vivo* closed-field acquisition to eliminate ambient light and equalize detection settings (e.g. imaging gains) [22], from whole resected tissue specimens to tumor cross sections to remove overlaying tissue in the imaging path [23] and minimizing difference in tumor size [4], from macroscopic to microscopic scale to take into account tumor cell density and delivery barrier [24] when characterizing molecular target and probe uptake. The interplay of these factors, among others (**Supplementary Table S4**), contributed to the translation of cellular expression of EGFR in individual tumor cells into NIR fluorescence TBR, which has implications for identification of molecular targets and appropriate tumor qualities for imaging. Our results have shown that the preferential breakdown of intratumoral delivery barrier such as in HGG can increase the specificity of fluorescence for tumor detection despite higher fluorescence heterogeneity. While total EGFR expression and delivery barrier correlated with overall panitumumab-IRDye800 distribution in tissue, tumor depth and operating room lighting are greater forces that ultimately determined the intraoperative tumor contrast at specific tissue surface locations of a certain patient. To accommodate a wide intraoperative signal range, minimal ambient light and the lowest imaging gain that allow tumor detection should be adopted to facilitate adoption and standardization of intraoperative protocols across institutions. This was an extension of our prior studies to characterize open-field imaging contrast with panitumumab-IRDye800 phantoms [25, 26].

We mapped a more comprehensive relationship between fluorescence contrast and margin distance in resected tissue specimens, while previous studies prioritized selection of a few high-risk NIR signal peaks for pathological assessment in head and neck cancer [23, 27, 28]. We further correlated NIR signals measured between two closed-field imaging devices to allow conversion of fluorescence measurements between devices and studies. Moreover, panitumumab-IRDye800 distribution registered to histology features at microscopic resolution (i.e. 21 μm, or 2 picograms of tissue) on intact tissue sections was characterized. Apart from intrinsic intratumoral and inter-patient heterogeneity across cancer types, fluorescence intensity of EGFR molecular targeting is a multi-factorial characteristic and may explain the wide range of reported EGFR levels in literature, as elucidated by our target validation results. Overall, our study is the first attempt to hold imaging performance of a particular optical probe intended for clinical translation in multiple cancers against a common acquisition and analytical framework so that clinicians and researchers can objectively evaluate its relative merits for different indications.

Among 20 cancer types in the Human Protein Pathology Atlas, moderate to strong EGFR positivity by immunohistochemistry was reported in 75% of malignant glioma and head and neck cancer patients, followed by 64% of lung cancer patients [29]. In this study, higher total tumoral EGFR expression, defined as percentage of area with positive immunoreactivity, correlated to greater panitumumab-IRDye800 concentration, fluorescence and TBR. However, HGG had the highest total EGFR expression (89%, followed by HNSCC: 62% and LAC: 41%) yet received less than half the panitumumab-IRDye800 delivery observed in HNSCC (3.9 vs. 8.1 ng/mg), suggesting delivery barrier. Conversely, intact blood-brain barrier (BBB) in normal tissue may result in improved tumor contrast during open-field imaging. Tight junction protein expression (Claudin-5, endothelial membrane) around blood vessels (ERG, endothelial nucleus) was used to assess the integrity of BBB, which was reduced in viable HGG tumor tissue compared to normal brain. Accordingly, specific cellular fluorescence in HGG tissue confirmed panitumumab-IRDye800 delivery across leaky blood-tumor barrier. This agreed with previous clinical [4, 30, 31] and preclinical [32] evidence that even modest EGFR expression was sufficient for tumor detection with panitumumab-IRDye800.

To mitigate confounding factors such as overlaying tissue and variable tissue thickness, intratumoral fluorescence contrast and cellular antibody delivery was measured on 4-μm tissue sections. In these optically transparent thin tissue sections, difference in light scattering properties among tumor types was negligible [33]. Although all the patients received a flat dose of 50 mg, due to the dose-dependent nature of panitumumab-IRDye800 half-life (14.5 h – 24.8 h in the 0.06 – 1.5 mg/kg dose range [2]) in patients, differences in body weight and imaging window can influence the plasma concentration of panitumumab-IRDye800 at the time of imaging and contribute to the variance in tissue fluorescence. Thus probe concentrations in individual tissue types were normalized by corresponding day-of-surgery plasma concentrations.

One of the limitations of the study is that tissue section thickness was subject to < 2% variability [34], and an average 11.4% tissue shrinkage during fixation and embedding [35] led to 12.9% overestimation of antibody concentration. To correct for these errors, tissue section fluorescence measurements were normalized by autofluorescence and the percentage of shrinkage. Since fluorescence is a surrogate for the presence of antibody in tissue, differences in dye-to-protein ratio across production batches, photo bleaching and metabolism can introduce noise and bias in any fluorescence-based method. Thus, direct antibody quantification techniques such as multiple reaction monitoring are being developed to validate and calibrate results from the fluorescence-based approach. Optimal dose finding was not included in this study, but rather has been reported separately. Nor were fluorescence used for intraoperative decision making per IRB protocols. Representation of some populations was lacking in these early phase trials.

We compared intraoperative and perioperative fluorescence imaging performance of a NIR-labeled EGFR antibody, panitumumab-IRDye800, infused in patients with HGG, HNSCC or LAC tumors. Antibody distribution correlated with EGFR expression, tumor cell density and biological factors. The analytical framework encompassing three cancer types and different imaging instruments offered a holistic perspective to collectively interpret the potential clinical utilities of molecular targeted fluorescence imaging in terms of tumor contrast enhancement, whole specimen margin assessment and antibody distribution in tissue, with implication in oncologically sound resections, informed decision-making on therapy and regulatory approval of new imaging probes that has the potential to transform standard-of-care practice and patient care.

## Methods

### Patients

This study was approved by the Stanford University Institutional Review Board (protocol numbers: 43179, 35064, and 41302). Patients (n = 31) undergoing surgery between Aug 2017 and Nov 2019 for high-grade glioma (HGG, n = 5), head and neck squamous cell carcinoma (HNSCC, n = 23) and lung adenocarcinoma (LAC, n = 3) were recruited in three clinical trials (NCT03510208, NCT02415881 and NCT03582124), respectively. Maximum dimension of tumor size was determined by pre-surgical magnetic resonance imaging (MRI) or computed tomography. A single dose of 50 mg panitumumab-IRDye800 was infused in patients 1 – 5 days prior to surgery, with intraoperative blood samples collected to measure plasma concentration (NIR plate reader, Tecan). Adverse events categorized according to the National Cancer Institute Common Terminology Criteria (Version 4.03) were collected up to 30 days after infusion. Areas of viable tumor as well as normal tissue were outlined by board-certified pathologists on representative H&E staining of tissue sections.

### NIR fluorescence imaging

On the day of surgery, a laparoscope or a portable handheld camera attached to the SPY fluorescence imaging platform (excitation/emission: 766 / > 800 nm; Novadaq, Burnaby, BC, Canada) detected intraoperative NIR fluorescence of the tumor and wound bed. Fluorescence was not used to make standard-of-care resection decisions. Solid tumors were resected en bloc with negative margins while diffuse HGGs were removed in small pieces. *Ex vivo* fresh tissue was imaged in a NIR florescence imager (excitation/emission: 785 / > 820 nm, resolution = 85 µm; Pearl Trilogy Imaging System, LI-COR Biosciences, Lincoln, NE) without ambient light [23, 36]. Solid tumors were then fixed in 10% neutral buffered formalin and sectioned into 5 mm-thick serial cross sections. FFPE tissue blocks were cut into 4 µm-thick tissue sections for histological and IHC staining. Fluorescence images of both tissue blocks and sections were acquired in a flatbed scanner (excitation: 685 nm and 785 nm; emission: > 810 nm; resolution = 21 µm; Odyssey CLx Infrared Imaging System, LI-COR Biosciences). The distance from the tissue resection surface to solid tumor margin was measured on H&E sections.

### Imaging quantification

Tumor contrast was measured by the ratio of average pixel intensities (ImageJ, 1.53c, NIH) from five randomly selected circular regions of interest (ROIs, d = 20 pixels) inside tumor and surrounding normal areas in intraoperative white light and fluorescence images in grayscale. Fluorescence histograms (pixel intensity: 0 – 255; pixel count: 0 – 5000) for the entire tumor and peritumoral normal areas were plotted. High-intensity peaks in the fluorescence map of resected tissue were isolated as previously described [28]. Mean fluorescence intensity (MFI) was measured as total fluorescence signal divided by the pixel number within ROIs (Image Studio, v5.2, LI-COR Biosciences). MFI in normal tissue was measured in muscle or brain tissue with < 20% tumor cells. Tumor-to-background ratio (TBR) on fresh resected tissue surface denoted the ratio of MFIs in circular ROIs (d = 2 mm) over tumor versus those over normal tissue. TBR of tissue sections was the ratio of MFIs within outlined tumor versus uninvolved tissue. Fluorescence heterogeneity denoted the standard deviation of fluorescence signal normalized by MFI. MFIs of anatomic structures (circular ROIs, d = 200 μm) on tissue sections were measured.

### Tissue-mimicking phantoms

Serial dilutions of panitumumab-IRDye800 (0 – 10.0 g/mL) were respectively dissolved in 1% agarose (Life Technologies) and 1% intralipid (Sigma-Aldrich) at 45°C and poured into 200 µL cylindrical molds. Solidified phantoms were imaged in the operating room under three lighting conditions (SPY platform, gain: 2, 4 and 8; ambient lights: TL-D, 36W, Philips; room lights: A19, 100W, Osram; overhead lights: F528, 140W, Stryker). The ratio of MFIs between panitumumab-IRDye800 and saline containing phantoms measured imaging contrast. Phantom MFIs were measured with both closed-field devices. Panitumumab-IRDye800 concentrations and MFIs of 4 μm FFPE phantom sections were fitted by polynomial regression.

### Immunohistochemical analysis

To examine EGFR expression, automated IHC and hematoxylin counterstaining were performed on FFPE tissue sections after heat mediated antigen retrieval with Dako Autostainer (Agilent Technologies; primary antibody: RM-2111-RQ, prediluted, Thermo Fisher Scientific; secondary antibody: SM805, prediluted, Agilent Technologies) including positive and negative controls. Double IHC staining of Claudin-5 (1:500, 34-1600, Thermo Fisher Scientific) and ETS-related gene (ERG, 1:1000, EPR3864, Abcam, Cambridge, United Kingdom) was performed on HGG tissue. Immunoreactivity was visualized with diaminobenzidine (for EGFR and Claudin-5) and magenta (for ERG) chromogens (Dako EnVision, Glostrup, Denmark). Microscopic images were acquired in a whole slide scanner (NanoZoomer 2.0-HT; Hamamatsu Photonics, Hamamatsu City, Japan). The percentage of pixels with moderate to strong staining was quantified with a counting algorithm (Image Scope Software, Aperio Technologies) as previously described [37]. EGFR+ tumor cells within tumor outlines were counted with a MATLAB (version R2021a) algorithm.

### Statistical analysis

Group statistics were expressed as mean ± standard error of the mean unless specified otherwise. Patient characteristics were compared between cancer types using analysis of variance (ANOVA) and Pearson’s Chi-square tests as appropriate. Paired t-tests (two-tailed) were performed for group comparisons between tumor and normal tissues in each cancer type. One-way ANOVA was performed for group comparisons among trials. Significance was defined at \**P* < 0.05; ** *P* < 0.01; *** *P* < 0.001; **** *P* < 0.0001. Whiskers and outliers of box plots were determined by the Tukey method. Receiver operating characteristics (ROC) curves were plotted for distinguishing histological tumor versus normal tissue using MFI and EGFR%, respectively. Sensitivity, specificity, area under the ROC curve (AUC), negative and positive predictive values (NPV & PPV) were subsequently calculated using these definitions. MFI and EGFR% cutoff values that resulted in the maximal sensitivity and specificity combined were reported.

## Data Availability

All data produced in the present study are available upon reasonable request to the authors.

## Acknowledgements

This work was supported in part by the Stanford Comprehensive Cancer Center, the Stanford University School of Medicine Medical Scholars Program, the Netherlands Organization for Scientific Research (Rubicon; 019.171LW.022), the National Institutes of Health and the National Cancer Institute (R01CA190306), National Institute on Deafness and Other Communication Disorders (T32DC015209), a scientific research grant from the Yokoyama Foundation for Clinical Pharmacology (YRY-1702), and an institutional equipment loan from LI-COR Biosciences Inc. Illustrations in Fig. 1, Fig. 4 and Fig. 5 were created by BioRender.com.

## Author Contributions

Conception and design: Q.Z. and E.L.R.

Development of methodology: Q.Z., N.S.vdB, W.K., N.N., M.A.E. S.vK. and E.L.R.

Review of the electronic medical records: Q.Z., N.S.vdB and M.A.E.

Acquisition of data: Q.Z., N.S.vdB, S.vK., M.A.E., J.C.M.V.L., Z.H., N.L., M.HG., G.L. and E.L.R.

Analysis and interpretation of data: all authors

Writing, review and/or revision of the manuscript: all authors

Administrative, technical, or material support: N.S.vdB., R.C.R., G.Y., M.G.

Study supervision: E.L.R.

Final approval of the version to be published: all authors

