## supplementary information for "Applications of fluorescence-guided surgery across multiple tumor types using a near-infrared labeled EGFR antibody"

**Supplementary Table S1.** Number of adverse events recorded within 30 days of panitumumab-IRDye800 infusion in three cancer types.

| | **HGG** (n = 5) | **HNSCC** (n = 23) | **LAC** (n = 3) | **Total** (n = 31) |
|---|---|---|---|---|
| Infusion reactions | No | No | No | No |
| Total serious adverse events | 0 | 0 | 0 | 0 |
| Total non-serious adverse events | 8 | 46 | 4 | 58 |
| (mean ± SD) | (1.75 ± 2.22) | (2.09 ± 1.85) | (1.33 ± 1.15) | (1.97 ± 1.80) |
| Grade I | 6 | 35 | 3 | 44 |
| Grade II | 2 | 8 | 1 | 11 |
| Grade III | 0 | 3 | 0 | 3 |
| Attribution | Unrelated | Unrelated | Unrelated | Unrelated |
| Dose limiting toxicity | No | No | No | No |

**Supplementary Table S2.** Diagnostic performance characteristics of tissue section NIR fluorescence for tumor detection, including sensitivity (Sen.), specificity (Spe.), positive predictive value (PPV), negative predictive value (NPV) area under the curve (AUC), and MFI cutoff values for maximal sensitivity and specificity combined.

| | **HGG** | **HNSCC** | **LAC** |
|---|---|---|---|
| Sensitivity | 72% | 89% | 92% |
| Specificity | 96% | 74% | 67% |
| PPV | 91% | 77% | 71% |
| NPV | 85% | 87% | 91% |
| AUC | 0.85 | 0.89 | 0.87 |
| MFI cutoff | 0.55 | 0.74 | 0.98 |

**Supplementary Table S3.** EGFR expression as a diagnostic tool for tumor on whole tissue sections. Performance was compared among three cancer types in terms of sensitivity (Sen.), specificity (Spe.), positive predictive value (PPV), negative predictive value (NPV), area under the curve (AUC) and EGFR+% cutoff values for maximal sensitivity and specificity combined.

| | **HGG** | **HNSCC** | **LAC** |
|---|---|---|---|
| Sensitivity | 96% | 97% | 80% |
| Specificity | 80% | 86% | 83% |
| PPV | 93% | 87% | 80% |
| NPV | 89% | 97% | 83% |
| AUC | 0.94 | 0.96 | 0.82 |
| EGFR+% cutoff | 66.71 | 17.63 | 29.65 |

HGG: high-grade glioma; HNSCC: head-and-neck squamous cell carcinoma; LAC: lung adenocarcinoma; PPV: positive predictive value; NPV: negative predictive value; AUC: area under the curve; EGFR: epidermal growth factor receptor

**Supplementary Table S4.** Factors contributing to NIR imaging performance of tumor in three cancer types.

| **Factors** | **HGG** | **HNSCC** | **LAC** |
|---|---|---|---|
| Cellular EGFR expression | +++ | ++ | ++ |
| Total tumoral EGFR expression | +++ | ++ | ++ |
| AUC of EGFR as biomarker | +++ | +++ | ++ |
| EGFR+ Tumor cell density | +++ | +++ | ++ |
| NIR in normal tissue *ex vivo* | + | ++ | +++ |
| NIR in tumor tissue ex vivo | ++ | ++++ | +++ |
| NIR tumor contrast ex vivo | +++ | +++ | ++ |
| AUC of NIR for tumor detection | ++ | +++ | ++ |
| Overlaying tissue above tumor | -/+ | -/++ | -/++ |
| WL tumor contrast in vivo | - | - | + |
| NIR tumor contrast *in vivo* | +++ | +++ | ++ |
| NIR wound bed contrast *in vivo* | - | - | - |

**Supplementary Figure S1** Fresh resected tumor tissue size from three trials. *** $P = 0.0001$ by ANOVA.

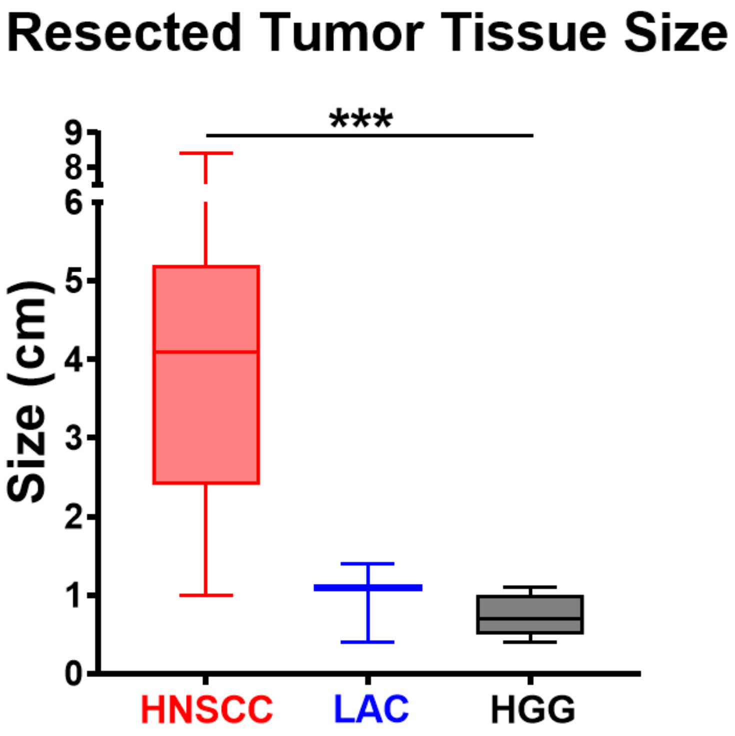

**Supplementary Figure S2** Neuronavigation identified the location (*asterisk*) of residual tumor in the wound bed on presurgical MR images, where language cortex involvement was indicated on fMRI mapping. P*ink*: visual responsive naming; *red*: object naming; *cyan*: auditory responsive naming; *blue*: negative BOLD signal; *grayscale*: preoperative T1+C; *glow*: T1 contrast enhancement.

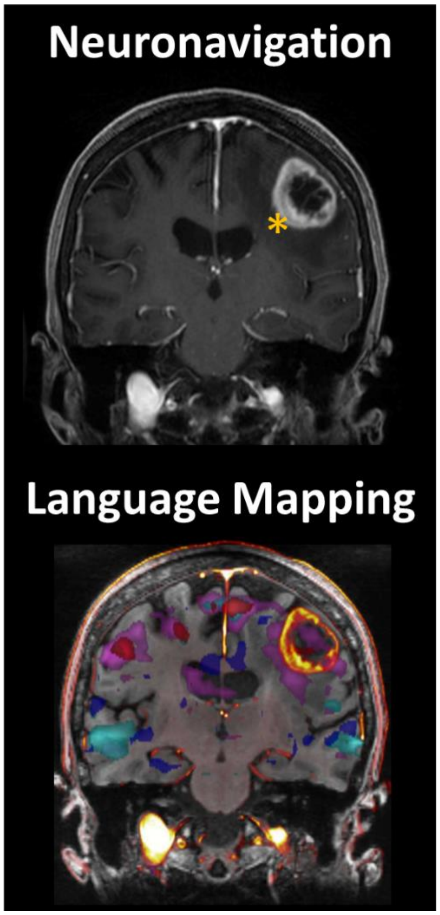

**Supplementary Figure S3** Correlation of fresh tumor tissue size with corresponding NIR target-to-background ratio (TBR). Each symbol represents one patient. Linear regression lines are fitted for each cancer type as well as for all patients combined.

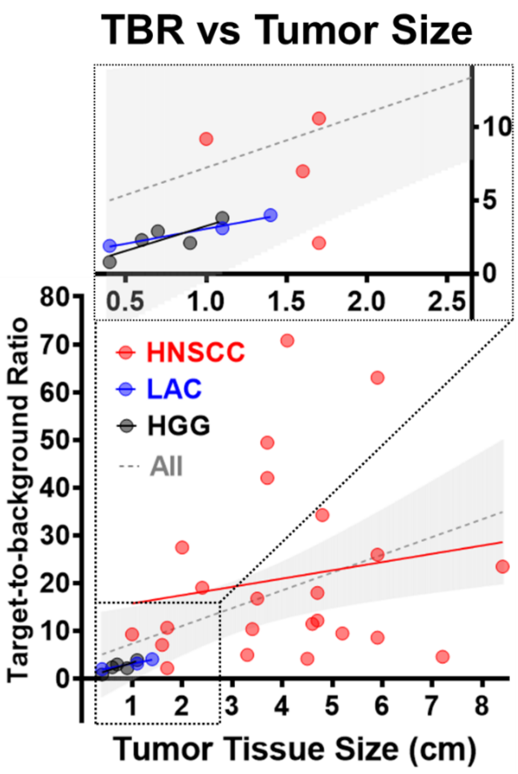

**Supplementary Figure S4** Open and closed-field NIR imaging with tissue mimicking phantoms. NIR fluorescence images of tissue-mimicking phantoms containing serial dilutions of panitumumab-IRDye800 (0 – 10 μg/mL) acquired in either an open-field imager under three lighting conditions with various gain settings, or a closed-field device. Open-field NIR imaging contrast (i.e. fluorescence intensity relative to that of a panitumumab-IRDye800 free phantom) of phantoms were plotted against the mean fluorescence intensity (MFI) of corresponding phantom imaged without ambient light (closed-field). The interquartile ranges of MFIs for fresh tumor tissue specimens (HGG, HNSCC and LAC) are indicated below the *X-axis*.

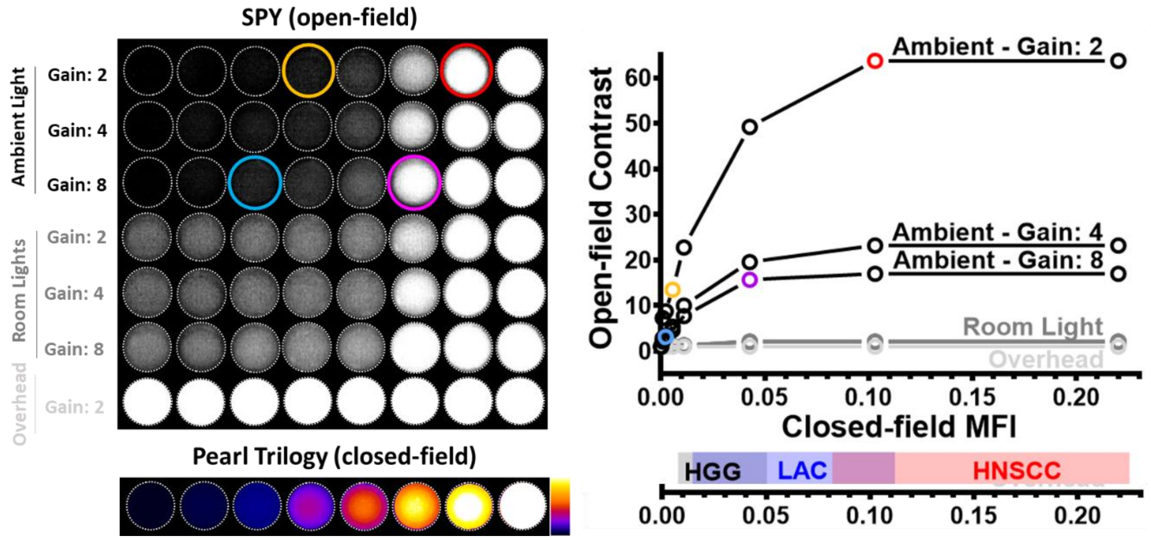

**Supplementary Figure S5** *Ex vivo* MFI by tissue type. Mean fluorescence intensity of fresh resected tumor (T) and normal (N) tissue (from left to right: $P < 0.0001$, $P = 0.2$, $P = 0.04$, $P = 0.08$), formalin fixed paraffin embedded tissue blocks (from left to right: $P < 0.0001$, $P < 0.0001$, $P = 0.25$, $P = 0.05$), and 4μm-thick tissue sections (from left to right: $P < 0.0001$, $P = 0.001$, $P = 0.6$, $P < 0.0001$). Paired t-test was performed for group comparisons.

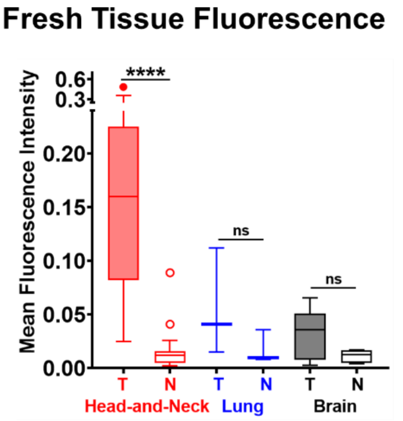


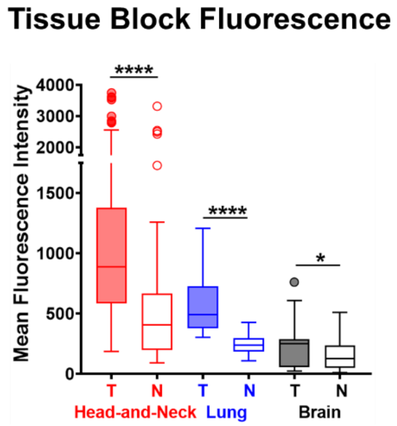


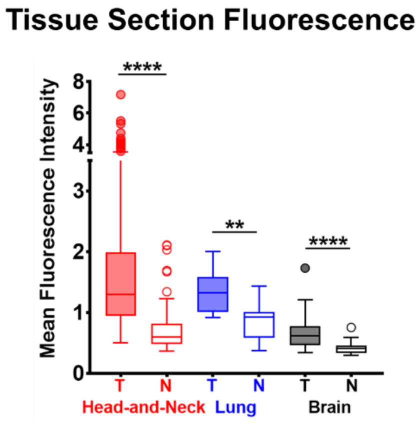

**Supplementary Figure S6** An array of phantoms imaged with two closed-field devices (*top*: Pearl Trilogy Imager; *bottom*: Odyssey CLx Flatbed Imager). Correlation of mean fluorescence intensities measured in each phantom between the two instruments. Each symbol is the average of three replicate measurements. Third order (cubic) polynomial least square curve fitting was performed.

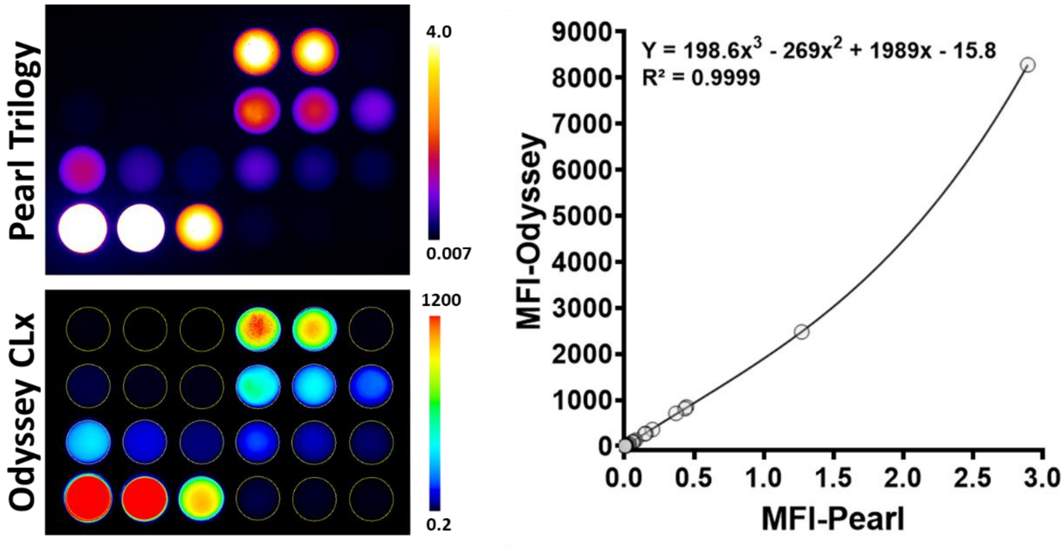

**Supplementary Figure S7** Standard curves of panitumumab-IRDye800 concentration versus mean fluorescence intensity in phantoms. Each symbol is the average of three replicate measurements. Third order (cubic) polynomial least square curve fitting were performed in the concentration ranges of 0 – 1 μg/mL and 1 – 10 μg/mL, respectively.

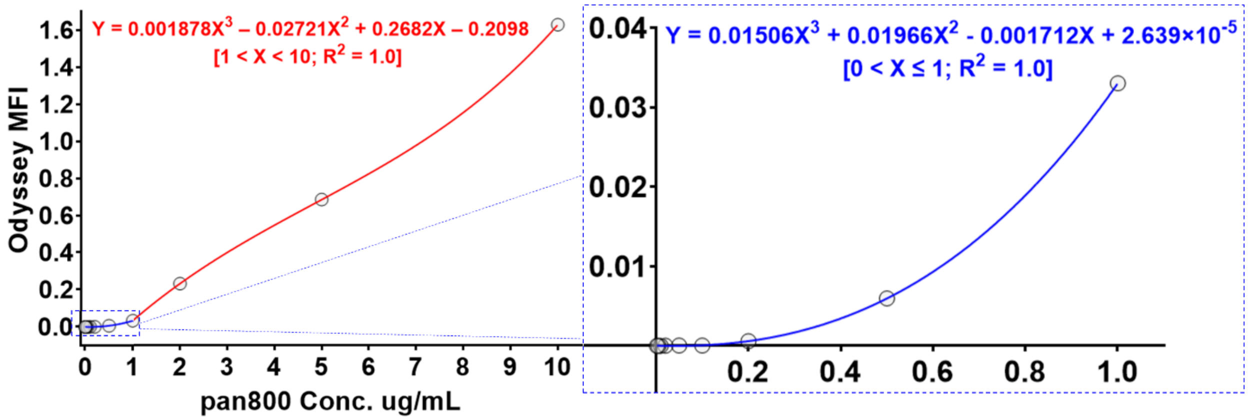

**Supplementary Figure S8** Double immunohistochemical staining of Claudin-5 (tight-junction protein, *brown*) and ETS-related gene (ERG, endothelial nucleus, *magenta*) on representative brain specimen containing normal brain and HGG tissue. *Arrows*: blood vessels; *dotted line*: infiltration edge.

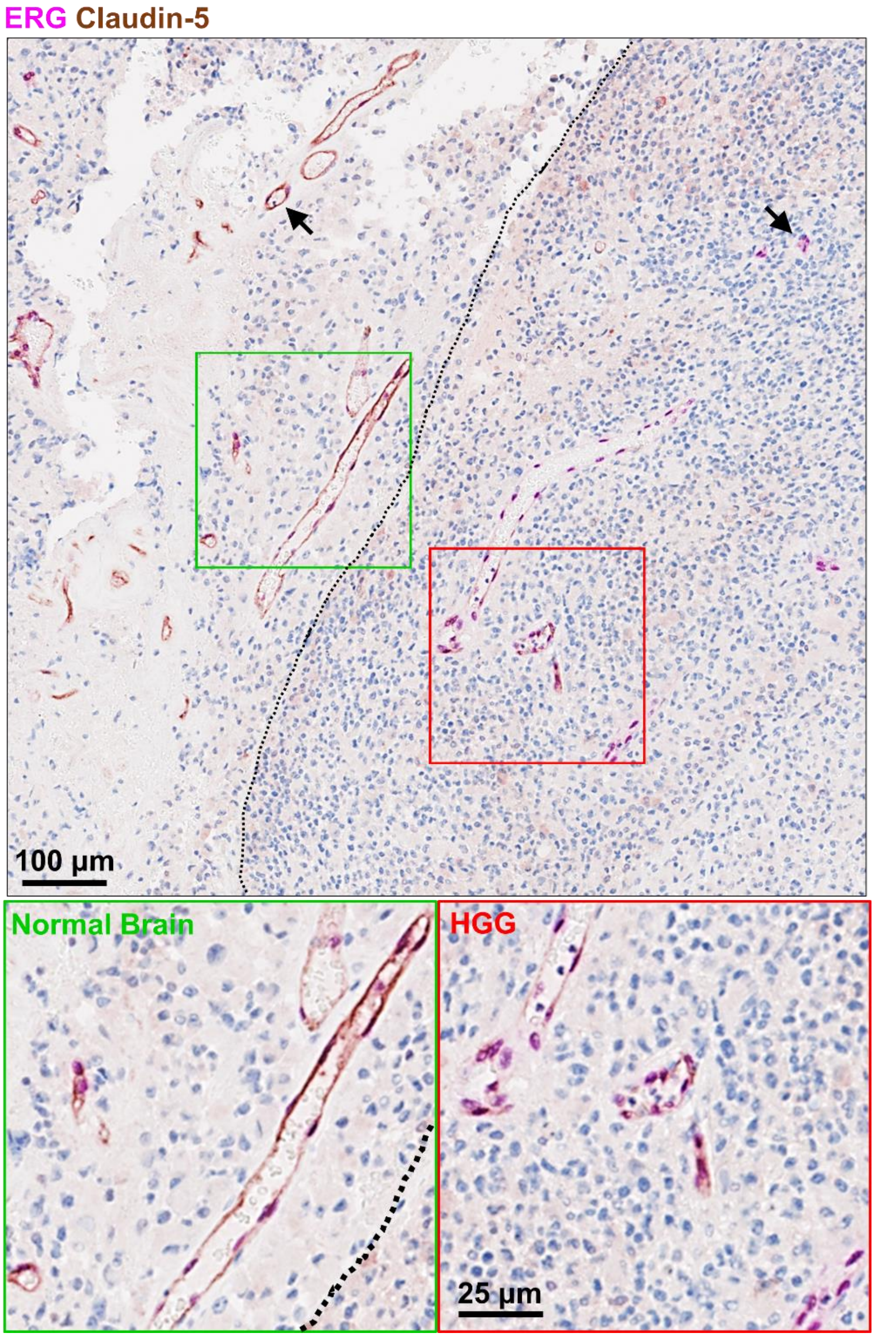

**Supplementary Figure S9** Heterogeneity of tumor cell density in three cancer types. Histograms of percentage of tumor area occupied by a certain number of tumor cells per mm$^2$ in HGG, HNSCC and LAC.

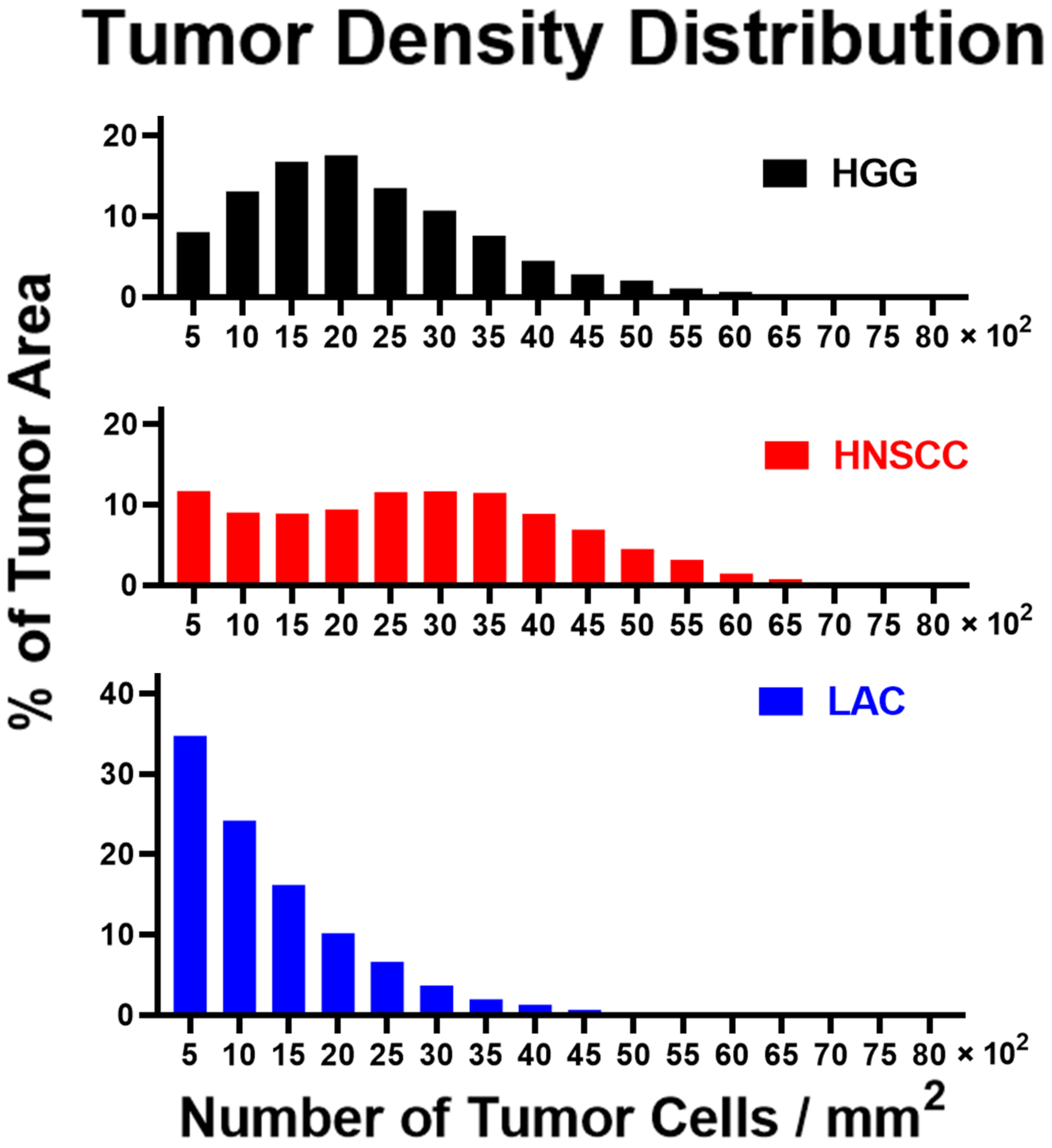

**Supplementary Figure S10** Immunohistochemical EGFR staining intensity mask. Immunohistochemical staining intensity maps of total tumoral EGFR, intratumoral EGFR and cellular EGFR expression in three cancer types. *Solid outlines*: tumor; *arrows*: areas of positive EGFR expression magnified in the 2nd column.

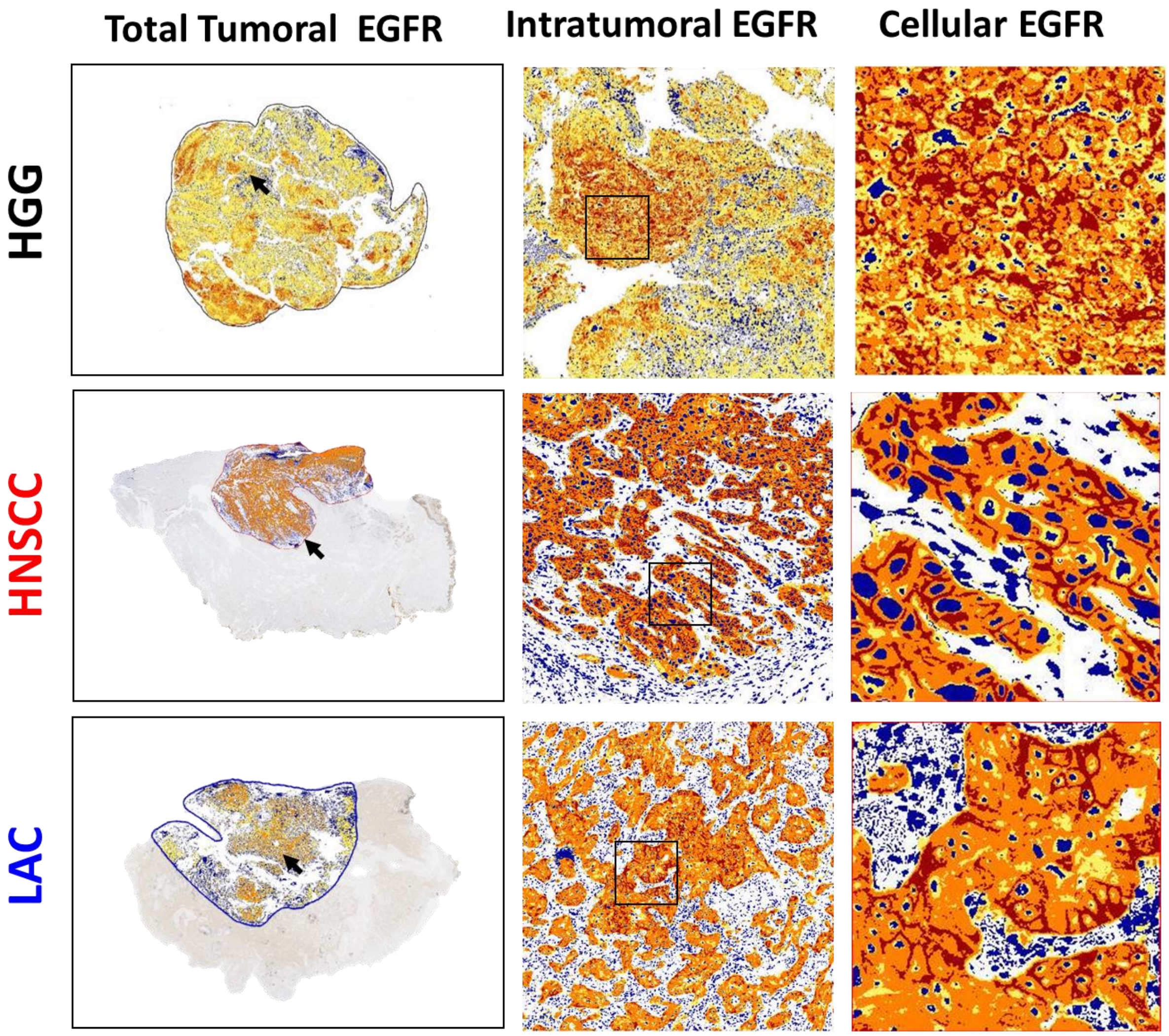
